# Modelling Lockdown Effects on Controlling the Spread of COVID-19

**DOI:** 10.1101/2022.06.04.22275993

**Authors:** W.K. Chow, C.L. Chow

## Abstract

The COVID-19 variant omicron is spreading rapidly, infecting about 1.2 million people in 2 months in Hong Kong from early January 2022. Locking down the city at the beginning of outbreak for an adequate period is effective in mitigation and suppression of disease transmission. However, it is difficult to implement the locking down proposal without strong supporting argument because of the accompanied economic loss. An appropriate mathematical model to provide key predictive information on the local epidemic and evaluate the effects of lockdown is reported in this paper. The number of susceptible citizens, infection cases and recovery number under some assumption on isolating citizens are predicted by solving the ordinary differential equations analytically. Observed infected cases during the fifth wave of outbreak in Hong Kong is taken as an example to illustrate the concept. Three lockdown scenarios are proposed and assessed by the developed mathematical approach. Early lockdown is illustrated to keep infected cases low, therefore very effective in controlling the spread by isolating the citizens in their own units.

## 1. Introduction

The spread of COVID-19 is rapid [1] with billions of people infected all over the world. The 5th wave of outbreaks in Hong Kong due to the variant Omicron [2,3] started in early January, 2022 after keeping zero daily infection cases for 7 months before that. The number of confirmed cases rose from over 2,000 on 14 February 2022, 34,000 on 8 March 2022, over 50,000 on 9 March 2022, then dropped to around 6,000 on 31 March 2022, 600 on 22 April 2022 and less than 300 in early May. Over 1.2 million citizens were infected. Hundred thousands of people were asked to stay at home due to limited capacity in designated healthcare centres.

Early lockdown of a city for a certain period might be effective in controlling the epidemic as demonstrated in many places. Objectives of lockdown are explained [4] to be for mitigation and suppression. Locking down will also avoid having too many infected cases in overloading the healthcare system. The healthcare system was overloaded in Hong Kong in early March 2022 [5], about one month after the outbreak. However, there are huge economic impact and it is always criticized by different stakeholders, particularly the servicing industry, including hotel and catering professionals. Proposal of locking down the city is difficult to be accepted without strong evidence. Further, there are problems on managing blocking procedure, distributing resources and medicines. The timing of lockdown is also important. Locking down too early when the number of infected cases is low can only be accepted in some places. But locking down late when many citizens are infected would be useless. It is impossible for the healthcare system to handle millions of infected citizens as experienced in Hong Kong.

Mathematical models are needed for predicting disease spread in time and the effects of proposed control scheme [6,7]. There were some predictions in Hong Kong from some mathematical models [8-13] in these months. Results predicted in early February 2022 indicated that hundred thousands of people would be infected in Hong Kong by March, 2022. Another one suggested that peak infection would be passed in March 2022. Predicted daily cases might drop below 1000 in mid-April, and less than 100 in May 2022. There were also estimation that over 4 million people had been infected up to April 2022. Up to 4 million people would be infected after starting the scheme in May 2022. All these predictions or estimations need to be justified before citizens trust and follow the control scheme.

Virus transmission is very complicated with many scenarios and scientific concepts behind in developing a deterministic model. Modelling particle trajectory only [14] needs numerous effort on simulating the basic physical phenomena. Only stochastic mathematical models [15-17] are available at the moment. However, the modelling parameters have to be determined or estimated from other similar cases through long-term observations.

Further, stochastic models available are good only for high population number. This is because the fundamental postulate [18] on the probability of an infection in the next period of time is proportional to the number of infectious individuals multiplied by the number of susceptible individuals. The number of infection cases is initially small (say less than 100) compared with the population size (7 million in Hong Kong). Short-term results predicted from the models should be justified [19,20] at the initial stage of the infection. Appropriate mathematical models studying transient effects within a few days after starting infection are needed.

Another key point is on the isolation time which depends on the time for virus taking effect and recovery time, depending on when the person is vaccinated. If the Omicron variant took about 5 days, the recovery criterion for those vaccinated 3 dosages is 7 days. Locking down for 7 days might be adequate. Others will be 14 days. Since over 1.1 million people were infected in 2 months, there were about 5 cycles. The number of people infected at a certain time during the 5^th^ wave is roughly 1,000,000/5 or 200,000, about 3% of 7 million population. This is quite consistent to the positive rate of 4% on testing.

Keeping infected persons in healthcare system or inside their units would cut the transmission chains effectively. A mathematical modelling approach on including the isolation effect on locking down the city is important and proposed to be studied in this paper.

## 2. Common Mathematical Modelling Approaches

The virus transmission mechanism through air is very complicated and hence it is difficult to develop deterministic models [21]. As reviewed [6], mathematical modelling is the only available method of predicting the extent of an emerging disease and assessing proposed control measures. There is little or no available data on previous outbreaks. Only stochastic models capture the inherent randomness in disease transmission observed in real-life outbreaks.

The basic model developed based on the number of susceptibility S, infection I and recovery R for a directly transmitted infectious disease is known as the SIR model as reviewed by [22]. In a discrete time model, the probability of an infection in the next period of time was proposed early [18] by Hamer in 1906 to be proportional to the number of infectious individuals multiplied by the number of susceptible individuals. That means faster the rate of lost 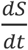 for larger S and I, i.e.

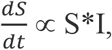

or

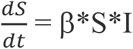

This assumption is acceptable for studying long-term infection with millions of people in months. However, for short-term studies in a week with less than 10,000 people infected out of a city of 7 million people, further justification is needed.

The SIR model has three coupled non-linear ordinary differential equations in S, I and R:

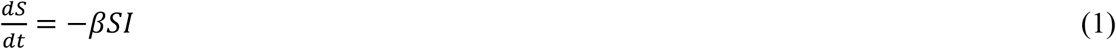

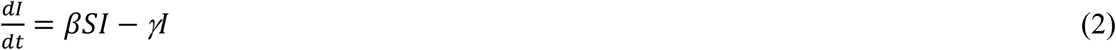

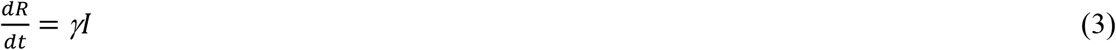

There is no explicit formula solution in general.

The key parameters involved above are the disease transmission rate β, recovery rate γ and duration of infection D, γ is related to D as:

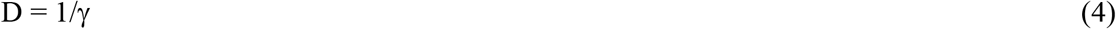

Summing up instantaneous values (say daily value) S(t), I(t) and R(t) gives the total number of population N_o_ as:

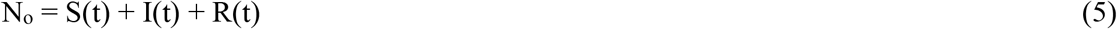

With this constraint, only two of the above three ordinary differential equations (1) to (3) need to be solved.

There are many other alternative forms of the above SIR type of model. A detailed disease-class transition dynamics was provided [23] from S to exposure E then to infectious I and removed I categories by using Erlang differential equation models of epidemic processes. This is also known as SEIR model. A mathematical model of transmission with four datasets from within and outside Wuhan was developed [19].

## 3. Review on Lockdown Effects

To avoid having rapid increase in infected cases, locking down is one possible way out. As reported, locking down would have two effects on mitigation and suppression [4]. Isolating infected persons would cut the transmission chains effectively. This scheme might be effective but early planning is required to handle management problems. Two points should be considered in lockdown:

- Number of infected cases Locking down early with low number of infected cases is effective but has economic impact. But locking down late would be many citizens infected, already overloaded the healthcare system as observed [5].
- Isolation If the Omicron variant took action about 5 days, the recovery time criterion for those vaccinated with 3 dosages is quarantined for 7 days, others without vaccination will be isolated for 14 days. Locking down for 14 days might be adequate for all people.

Epidemic models able to predict the future number of active cases and deaths when lockdowns with different stringency levels or durations are needed. As reported, the key observation is that lockdown-induced modifications may not be captured by a traditional compartmental models because these models assume uniformity of social interactions among the population [24]. A model [24] able to capture the abrupt social habit changes caused by lockdowns was reported. Another mathematical model of COVID-19 dynamics was developed to include age-stratified disease parameters and contact matrices [25]. Different lockdown scenarios were simulated. Results suggested that reducing work contacts is more efficient than reducing school contacts, or implementing shielding for people over 60. The dynamic behaviour of COVID-19 spreading after a lockdown lifting [26] was studied.

The SIR model itself has not yet included the above isolation effects. Including exposure E to give a SEIR model might be more appropriate to study locking down effects. Exposed people member would only affect people staying inside the same unit with them. Units without infected members got zero E. A modified SEIR model [27] was proposed by adding another equation on containment.

## 4. Analytical Study Proposed

The SIR model given by equations (1) to (3) is useful to study the isolation effect. Applying isolation with the infection number *I*_*c*_, corresponding *S*_*c*_ and *R*_*c*_, at time *t*_*c*_, satisfying

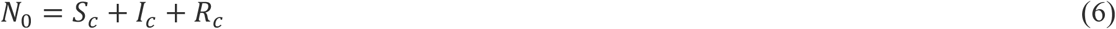

Suppose taking action early so that people can be isolated in separation units. Individual isolation of the infected means *I* will be kept at value *I*_*c*_,

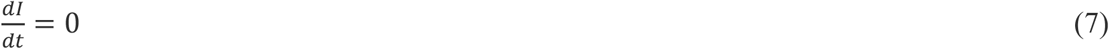

From time *t* > *t*_*c*,_ equations on *S, I* and *R* become:

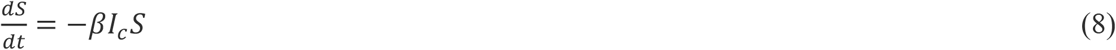

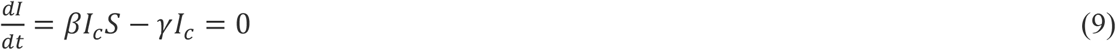

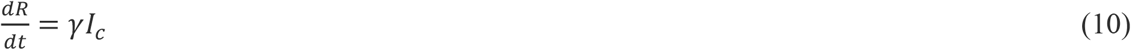

Integrating equation (8) gives

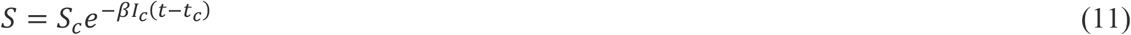

*S* will be kept at *S*_*c*_ for single person isolation. Integrating equation (10) gives

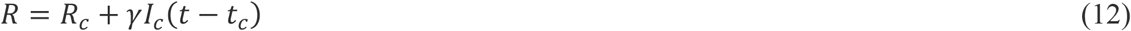

After time interval *τ, I* starts to decay at *t*. At this stage,

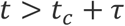

Equation for *I* is:

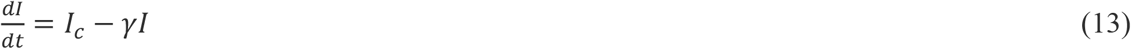

Integrating equation (13) gives

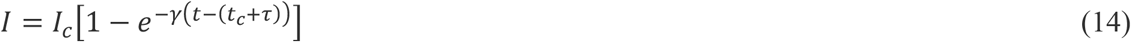

*R* equation is the same as (3), putting in equation (14) gives

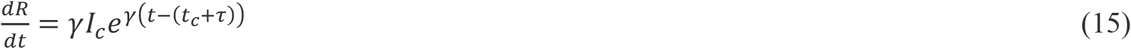

or

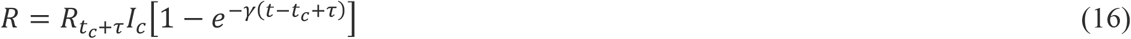

Suppose locking down actions take time at *t*_*c*_ when I = I_c_, S = S_c_. The infected persons are isolated by not allowing them to leave their unit. If each unit has m_av_ persons, the maximum number of persons infected after containment is I_c_m_av_.

## 5. Three Locking Down Scenarios

The cumulative infection number I of the 5^th^ wave outbreak in Hong Kong plotted in Fig. 1 is taken as an example to study lockdown effect.

**Fig. 1:**
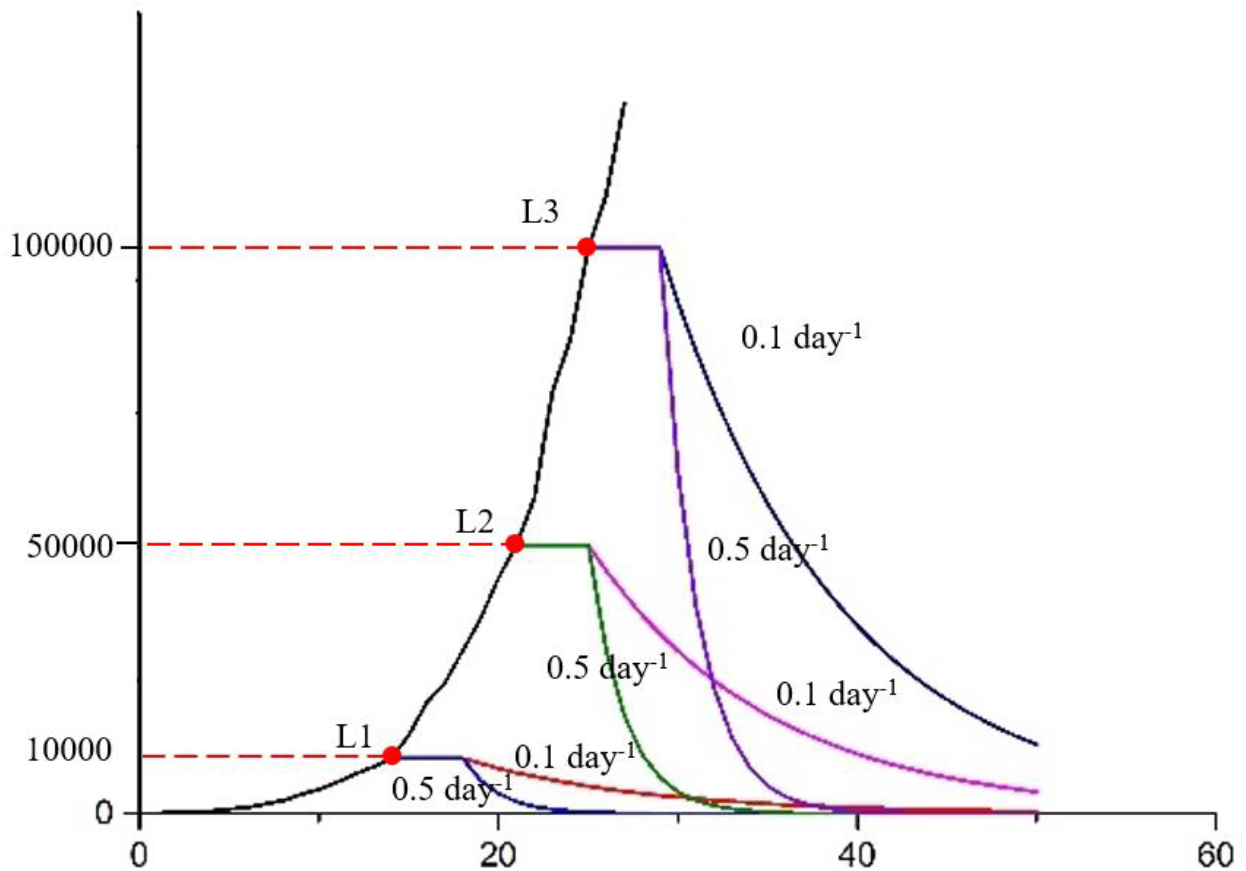
The three locking down scenarios

The city after having COVID outbreaks for a certain period, the infection duration D in equation (4) above can be estimated by observing the infection characteristics. Keeping all citizens in their own units for a period D, say 2 days to 10 days with an average of 5 days as the active period of Omicron variant, might be considered to cutoff the transmission chains. However, those family members staying with the infected persons might be affected too. Infected people with symptoms can be picked up readily because they need medical care. However, those infected persons with no symptoms cannot be identified easily without carrying out a territory-wide compulsory testing.

Three locking down scenarios are proposed and considered in this study for γ in equations (2) and (3) taken as 0.1 day^−1^ and 0.5 day^−1^:

- Scenario L1: Suppose isolation starts when the number of infected cases gives up to 10,000.
- Scenario L2: Arrange isolation later when *I* is 50,000. Results are plotted in Fig. 1 on *I*. Isolating infected people in separate units by themselves would stop the virus transmission in view of Fig. 1 on estimation I from equation (14).
- Scenario L3: Isolating at a later stage when 100,000 infected.

As observed from Fig. 1, blocking all 10,000 people in L1 single by themselves would stop transmission from infected carrier. The infected number will stay at constant once blocking starts. I then dropped after time at a rate depends on γ. But this is not viable as it is difficult to keep a single person by themselves.

Suppose a unit has an average of m_av_ people, of say 5, blocking each unit will have a maximum of 5 I_c_ or 50,000, the scenario then becomes L2. Infected number kept at 50,000, and then decreases after some time as in Fig. 1. Average number of persons m_av_ as 5, infecting 10,000 persons in Scenario L1 means a maximum of 50,000 will be infected. The infection curve is similar to Scenario L2 as shown in Fig. 1, and those 50,000 persons will not affect others. If they have symptoms, particularly those without vaccination, medical care is needed and they will contact healthcare systems by themselves. The transmission chains will also be cut, but the healthcare systems might be overloaded as in Hong Kong during the 5th wave outbreaks.

Isolating at a later stage when I_c_ is at 100,000 for L3 will have much more infected persons as in Fig. 1 even with individual isolation. A maximum of 500,000 citizens will be infected and then recovered with I decreased.

The increasing curve as in Fig 1, rose to 1.1 million within 2 months was observed in Hong Kong. Shenzhen took quick lockdown actions at early stage of outbreaks for a week. This is similar to Scenario L1 as the lower curve plotted in Fig 1. The number of confirmed infected cases was very limited, and the city resumed normal operation a week. Situation at Shanghai is in the middle of Fig. 1, infected over 100,000 already in a week, daily infected around 20,000 since 9 April, 2022.

Identification tests on virus are complicated [28-32]. It is difficult to implement such tests in places without adequate testing laboratory facilities on polymerase chain reaction (PCR) tests. Accuracy of the rapid antigen tests (RAT) is not high, particularly for the elderly without training on tests nor using smart phones. The supporting management practice of arranging such mass scale testing, such as on food and regular medicine, is another concern for big cities. Effectively, asymptomatic carriers are very difficult to identify in many places.

Two points on isolation and exposure are worthwhile to consider. Isolation of people inside their units would slow down the spread of the most vulnerable population, particularly to the elderly. Exposure to young healthy persons to virus would develop immune. But this is very dangerous to the elderly. But isolating people in the units can limit the infection within the units, not to the elderly outside.

## 6. Conclusion

A modeling approach on isolation effect is developed to study transient effects of the number of infection cases within a short time as reported in this paper. The assumptions made are explained on above. Analytical simulation of the ordinary differential equations on number of susceptible infection cases of recovery were achieved. Three lockdown scenarios are proposed and assessed as discussed above.

To control outbreak of COVID-19, locking down the city at the beginning stage for an adequate period is effective in mitigation and suppression as illustrated by the above study. It might be easier to consider the locking down proposal with predictions using the model developed above. An emergency planning to support residents while locking down.

With 10% of citizens infected, territory-wide compulsory universal COVID testing and locking down the city for a period matching with the characteristics of various variants might be necessary for mitigation and suppression of the transmission. Compulsory testing for all citizens would pick up asymptomatic carriers. Locking down the city by keeping people staying at designated accommodations, or at their own unit for mild or even no symptoms for those vaccinated would cut off or at least delay further infection.

## Data Availability

All data produced in the present study are available upon reasonable request to the authors

